# Accidental Exposure to Body Fluids Among Healthcare Workers in a Referral Hospital in the Security-Challenged Region of South West Cameroon

**DOI:** 10.1101/2024.02.20.24303093

**Authors:** Innocent Takougang, Fabrice Zobel Lekeumo Cheuyem, Blessing Asongu Changeh, Ngati Denetria Nyonga, Hortense Mengong Moneboulou

## Abstract

**Introduction:** Accidental exposure to body fluids (AEBs) increases the risk of blood-borne infections among susceptible HCWs. While 90% of the AEB reported occur in developed nations, developing countries bear 90% of the burden of healthcare associated infections, especially those of sub-Saharan Africa. Social insecurity may contribute further to the vulnerability of HCWs. Our study sought to determine the prevalence, reporting and management of AEBs among HCWs in the security-challenged Region of South-West Cameroon.

**Methods:** A cross-sectional study was carried out from February 2023 to April 2023, at the Buea Regional Hospital. Following informed consent, a 28-item interviewer-administered questionnaire to HCW was used. Data was entered and analyzed using R Statistics version 4.3.1.

**Results:** Out of the 230 HCWs that were approached, 200 were responded for a participation rate of >85%. The prevalence of AEB was high (93%). Exposures occurred while administering injections (37%), during blood sample collection (16%), delivery (11%), surgery (10.2%) and washing. The main risk factors for AEB included female gender (aOR=2.86) and those exercising in the medical (aOR=5.95), pediatrics (aOR=10.5), obstetrical (aOR=22.6), dental (aOR=26.3) units. Only 46.8% of AEBs were reported. Post-exposure management was carried out for 67.2% of the reported cases. Most HCW were unaware of the existence of an Infection Control Committee within the study setting, corroborating gaps in the observance of Standard Precautions.

**Conclusions:** Most HCWs experienced AEBs over the last year. There is a need to sensitize and enforce the observance of universal precautions among HCW of the Buea Regional Hospital. Such measures should be extended to other health facilities in related settings.

## Background

Healthcare associated infections (HAIs) are contracted during a stay or visit in a hospital environment, as a result of an unintended contact with blood or other body fluids (BBFs)[1–4]. Factors favoring accidental exposure to BBFs among healthcare workers (HCWs) include host and institutional determinants [4]. Such exposures are a burden to society increasing the costs of treatment, absence from work, distress, anxiety and early retirement [5–8]. Their financial burden and mortality are related to the virulence of the pathogenic agents involved and their antibiotic resistance [9–12]. Although the phenomenon of HAI is global, Africa is disproportionately affected [13–16]. HAIs result from failures in the implementation of hygiene and adequate infection transmission precautions in healthcare facilities. Such measures are essential in the promotion and delivery of quality healthcare [15]. More than 3 million HCWs million healthcare workers experience an accidental needle stick and sharp injuries (NSIs) every year, thus getting exposed to as many as twenty blood-borne pathogens, resulting in 170,000 to HIV infections, 2 million to HBV infections and 0.9 million to HCV infections [18,19]. While 90% of accidental exposures occur in the developing world, more than 90% of infections occur in developing countries, particularly in Africa where the prevalence of the underlying infections is high and adherence to standard precautions are poor [18,19]. Such situations are exacerbated by conflicts, insecurity and social unrest [20]. Given their socio-economic burden, some countries have established surveillance systems to manage exposures to blood-borne infections in healthcare settings [21]. In many African countries, such systems are plagued by underreporting and poor documentation [22–24]. The lack of training is a major hinderer to the observance of standard precautions [25]. Study reports shown that splashes were the most reported AEB and most victims worked in the surgical department [26,27]. Risk perceptions and trust in post-exposure management affected the reporting patterns among HCWs [28,29]. The implementation of Infection control and prevention measures (ICP) is typically coordinated by the Infection Control Committee (ICC) where operational data are used to steer interventions [31-33]. In the Cameroon healthcare pyramid, the Regional Hospitals (RH) typically offers surgery, internal medicine, obstetrics & gynecology, laboratory, pediatrics and emergency services; assuming capacities building roles for their satellite and district health facilities. Such roles include infection prevention and quality of healthcare services. Our study sought to investigate the determinants of Accidental Exposures to Blood and Body fluids among healthcare workers within the security-challenged Region of South-West Cameroon.

## Methods

We conducted a hospital-based cross-sectional study with descriptive and analytical aims from February to April 2023.

An exhaustive sample of HCWs of the Buea Regional Hospital were targeted for inclusion. The Regional Hospitals are the second level of reference. This facility has a capacity of 110 beds and a catchment population of over 200,000 inhabitants. The clinical services include surgery, emergency, hemodialysis, anesthesia-reanimation, ophthalmology, radiology, pediatric, cardiology, gynecology and obstetrics, internal medicine, odonto-stomatology, ear, nose and throat, laboratory, cleaning and sterilization departments. The study population included HCWs. These were nurses, laboratory technicians, midwives, assistant nurses, general practitioners, cleaners, dental surgeons and medical specialists, who had the potential to be exposed to BBFs in the execution of their duties and who gave their informed consent. A self-administered pre-tested 28 item-questionnaire was used, covering socio-professional characteristics, history of exposure, adherence to standard precautions, reporting and management of AEBs.

Following collection, data were entered, cross-checked and analyzed using R Statistics version 4.3.1. Simple and multiple binary logistic regressions were used to assess the strength of the association between variables. The selection of predictors that best fit the model was assessed using the stepwise Akaike Information Criterion (AIC). A *p*-value <0.05 was considered statistically significant. Confidence intervals (CI) were estimated at the 95% level.

The dependent variables included experience of exposure (NSIs and splashes) over the past 12 months and the reporting of the last exposure. Independent variables included age, sex, education level, socio-professional status and healthcare related practices.

The study protocol was approved by Institutional Review Board of the Faculty of Medicine and Biomedical Sciences – The University of Yaoundé I, under the ethical clearance: N°0344/UY1/FMSB/VDRC/DAASR/CSD. Informed consent was obtained from participants prior to inclusion in the study. All methods were performed in accordance with the declaration of Helsinki on research involving human subjects.

## Results

Out of 230 HCWs that were contacted, 200 completed and returned the questionnaires for a response rate of 87%.

### Occupational Exposure Experience

Over 83.5% (CI: 77.6-88.4%) of the study participants experienced an occupational exposure over the last 12 months. Exposure involved NSI (48.5%; CI: 41.4-55.7%) or splash (35.5%; CI: 28.9-42.6%).

### Factors Associated with Occupational Exposure to Body Fluids

Participants working in the internal medicine (aOR=5.95; *p*-value=0.006) and pediatric (aOR=10.9; *p*-value=0.008) units were more likely to experience NSI. The pattern of exposure revealed a gender difference as male HCWs were 2 times more like to experience a NSI (Table 1).

**Table 1.** Factors Associated with Needle Stick Injuries (NSI) Among Healthcare Workers of the Buea Regional Hospital, 2023 (*n*=200)

| Risk Factor | NSI<br>*n* (%) |  | Total<br>(100%) | cOR | *p*-value | aOR | 95%CI |  | *p*-value |
| --- | --- | --- | --- | --- | --- | --- | --- | --- | --- |
|  | Yes<br>*n*=97 | No<br>*n*=103 |  |  |  |  | Lower | Upper |  |
| **Sex** |  |  |  |  |  |  |  |  |  |
| Female | 69 (46.0) | 81 (54.0) | 150 | 1 |  | 1 | — | — |  |
| Male | 28 (56.0) | 22 (44.0) | 50 | 1.49 | 0.2218 | 2.05 | 1.01 | 4.40 | 0.0517 |
| **Age group (years)** |  |  |  |  |  |  |  |  |  |
| 18-24 | 12 (44.4) | 15 (55.6) | 27 | 1 |  |  |  |  |  |
| 25-34 | 60 (48.8) | 63 (51.2) | 123 | 1.19 | 0.6832 |  |  |  |  |
| 35-44 | 19 (50.0) | 19 (50.0) | 38 | 1.25 | 0.6587 |  |  |  |  |
| 45 + | 6 (50.0) | 6 (50.0) | 18 | 1.25 | 0.7482 |  |  |  |  |
| **Educational level** |  |  |  |  |  |  |  |  |  |
| University | 90 (52.9) | 80 (47.1) | 180 | 1 |  |  |  |  |  |
| Secondary or Primary School | 13 (43.3) | 17 (56.7) | 20 | 1.47 | 0.3335 |  |  |  |  |
| **Unit** |  |  |  |  |  |  |  |  |  |
| Laboratory | 4 (22.2) | 14 (77.8) | 18 | 1 |  | 1 | — | — |  |
| Medical ward | 37 (59.7) | 25 (40.3) | 62 | 5.18 | **0.0083** | 5.95 | 1.77 | 24.3 | **0.0066** |
| Surgical ward | 19 (48.7) | 20 (51.3) | 39 | 3.33 | 0.0650 | 3.74 | 1.06 | 15.7 | 0.0505 |
| Obstetrics and gynecology | 5 (22.7) | 17 (77.3) | 22 | 1.03 | 0.9696 | 1.07 | 0.23 | 5.29 | 0.9296 |
| Pediatrics | 9 (75.0) | 3 (25.0) | 12 | 10.5 | **0.0072** | 10.9 | 2.07 | 74.3 | **0.0079** |
| Dental service | 2 (33.3) | 4 (66.7) | 6 | 1.75 | 0.5888 | 1.27 | 0.13 | 10.1 | 0.8210 |
| Intensive care unit | 3 (50.0) | 3 (50.0) | 6 | 3.50 | 0.2076 | 4.22 | 0.56 | 34.5 | 0.1602 |
| Emergency | 9 (56.3) | 7 (43.8) | 16 | 4.50 | **0.0474** | 4.24 | 0.96 | 21.7 | 0.0657 |
| Out - patient consultation | 9 (47.4) | 10 (52.6) | 19 | 3.15 | 0.1159 | 2.87 | 0.69 | 13.6 | 0.1594 |
| **Professional status** |  |  |  |  |  |  |  |  |  |
| Laboratory technician | 4 (23.5%) | 13 (76.5%) | 17 | 1 |  |  |  |  |  |
| Physician | 11 (57.9%) | 8 (42.1%) | 19 | 4.47 | **0.0422** |  |  |  |  |
| Midwife & Nurse | 70 (48.6%) | 74 (51.4%) | 144 | 3.07 | 0.0593 |  |  |  |  |
| Assistant nurse & Cleaner | 12 (60.0%) | 8 (40.0%) | 20 | 4.88 | **0.0304** |  |  |  |  |
| **Years of experience 1** |  |  |  |  |  |  |  |  |  |
| <5 | 63 (48.1) | 68 (51.9) | 131 | 1 |  |  |  |  |  |
| 5+ | 34 (49.3) | 35 (50.7) | 69 | 1.05 | 0.8735 |  |  |  |  |
| **Years of experience 2** |  |  |  |  |  |  |  |  |  |
| <10 | 81 (48.5) | 86 (51.5) | 167 |  |  |  |  |  |  |
| 10+ | 16 (48.5) | 17 (51.5) | 33 | 1.00 | 0.9985 |  |  |  |  |
| **Glove always available** |  |  |  |  |  |  |  |  |  |
| Yes | 58 (46.0) | 68 (54.0) | 126 | 1 |  |  |  |  |  |
| No | 39 (52.7) | 35 (47.3) | 74 | 1.31 | 0.3625 |  |  |  |  |
| **Awareness of ICC** |  |  |  |  |  |  |  |  |  |
| Yes | 18 (32.1) | 38 (67.9) | 56 | 1 |  |  |  |  |  |
| No | 2 (40.0) | 3 (60.0) | 5 | 1.41 | 0.7209 |  |  |  |  |
| I don't know | 77 (55.4) | 62 (44.6) | 139 | 2.62 | **0.0038** |  |  |  |  |
| **Recapping needle practice** |  |  |  |  |  |  |  |  |  |
| No | 10 (38.5) | 16 (61.5) | 26 | 1 |  |  |  |  |  |
| Yes | 87 (50.0) | 87 (50.0) | 174 | 1.60 | 0.2751 |  |  |  |  |
| **ICP refresher training received** |  |  |  |  |  |  |  |  |  |
| Yes | 17 (35.4) | 31 (64.6) | 48 | 1 |  | 1 | — | — |  |
| No | 80 (52.6) | 72 (47.4) | 152 | 2.03 | **0.0394** | 2.06 | 1.01 | 4.30 | 0.0506 |
| **Poster on AEB available** |  |  |  |  |  |  |  |  |  |
| Yes | 88 (47.8) | 96 (52.2) | 184 | 1 |  |  |  |  |  |
| No | 7 (53.8) | 6 (46.2) | 13 | 1.27 | 0.6752 |  |  |  |  |
| I don't know | 2 (66.7) | 1 (33.3) | 3 | 2.18 | 0.5271 |  |  |  |  |

*cOR: Crude Odds Ratio; aOR: Adjusted Odds Ratio; CI: Confidence Interval, ICP: Infection Control and Prevention, ICC: Infection Control and Prevention Committee, AEB: Accidental Exposure to Body fluids; NSI: Needlestick and Sharp Injury*

Female HCWs were 2.9 times more likely to experience a splash mediated exposure to BBFs (*p*-value=0.019). The work was associated with splashes. Participants working in the laboratory (aOR=13.3; p-value=0.004), obstetric (aOR=23.3; *p*-value=0.0008) and dental (aOR=26.3; *p*-value=0.006) services experienced the most splash exposures (Table 2).

**Table 2.** Factors Associated with Splash Exposure Among Healthcare Workers at the Buea Regional Hospital, 2023 (*n*=200)

| Risk Factor | Splash Exposure<br>*n* (%) |  | Total<br>(100%) | cOR | *p*-value | aOR | 95%CI |  | *p*-value |
| --- | --- | --- | --- | --- | --- | --- | --- | --- | --- |
|  | Yes<br>*n*=71 | No<br>*n*=129 |  |  |  |  | Lower | Upper |  |
| **Sex** |  |  |  |  |  |  |  |  |  |
| Male | 13 (26.0) | 37 (74.0) | 50 | 1 |  | 1 | — | — |  |
| Female | 58 (38.7) | 92 (61.3) | 150 | 1.79 | 0.1077 | 2.86 | 1.23 | 7.18 | **0.0188** |
| **Age group (years)** |  |  |  |  |  |  |  |  |  |
| 18-24 | 11 (40.7) | 16 (59.3) | 27 | 1 |  |  |  |  |  |
| 25-34 | 42 (34.1) | 81 (65.9) | 123 | 0.75 | 0.5171 |  |  |  |  |
| 35-44 | 15 (39.5) | 23 (60.5) | 38 | 0.95 | 0.9182 |  |  |  |  |
| 45 + | 3 (25.0) | 9 (75.0) | 12 | 0.48 | 0.3491 |  |  |  |  |
| **Educational level** |  |  |  |  |  |  |  |  |  |
| University | 64 (37.6) | 106 (62.4) | 170 | 1 |  |  |  |  |  |
| Secondary or Primary School | 7 (23.3) | 23 (76.7) | 30 | 0.50 | 0.1365 | 0.35 | 0.011 | 1.01 | 0.0638 |
| **Unit/Service** |  |  |  |  |  |  |  |  |  |
| Emergency | 2 (12.5) | 14 (87.5) | 16 | 1 |  | 1 | — | — |  |
| Laboratory | 11 (61.1) | 7 (38.9) | 18 | 11.0 | **0.0075** | 13.3 | 2.51 | 109 | **0.0043** |
| Medical ward | 18 (29.0) | 44 (71.0) | 39 | 2.86 | 0.1918 | 2.58 | 0.61 | 18.0 | 0.2492 |
| Surgical ward | 9 (23.1) | 30 (76.9) | 22 | 2.10 | 0.3805 | 1.92 | 0.41 | 14.0 | 0.4493 |
| Obstetrics and gynecology | 17 (77.3) | 5 (22.7) | 12 | 23.8 | **0.0005** | 22.6 | 4.30 | 186 | **0.0008** |
| Pediatrics | 3 (25.0) | 9 (75.0) | 6 | 2.33 | 0.4005 | 1.66 | 0.21 | 15.5 | 0.6278 |
| Dental service | 4 (66.7) | 2 (33.3) | 6 | 14.0 | **0.0217** | 26.3 | 2.91 | 361 | **0.0063** |
| Intensive care unit | 0 (0.0) | 6 (100.0) | 6 | — |  | — | — | — | 0.9879 |
| Out - patient consultation | 7 (36.8) | 12 (63.2) | 19 | 4.08 | 0.1152 | 4.65 | 0.87 | 36.7 | 0.0937 |
| **Professional status** |  |  |  |  |  |  |  |  |  |
| Assistant nurse & Cleaner | 15 (75.0) | 5 (25.0) | 20 | 1 |  |  |  |  |  |
| Laboratory technician | 6 (35.3) | 11 (64.7) | 17 | 5.50 | **0.0185** |  |  |  |  |
| Physician | 12 (63.2) | 7 (36.8) | 19 | 1.75 | 0.4254 |  |  |  |  |
| Midwife & Nurse | 96 (66.7) | 48 (33.3) | 144 | 1.50 | 0.4576 |  |  |  |  |
| **Years of experience 1** |  |  |  |  |  |  |  |  |  |
| <5 | 46 (35.1) | 85 (64.9) | 131 | 1 |  |  |  |  |  |
| 5+ | 25 (36.2) | 44 (63.8) | 69 | 1.05 | 0.8753 |  |  |  |  |
| **Years of experience 2** |  |  |  |  |  |  |  |  |  |
| <10 | 58 (34.7) | 109 (65.3) | 167 | 1 |  | 1 | — | — |  |
| 10+ | 13 (39.4) | 20 (60.6) | 33 | 1.22 | 0.6093 | 2.08 | 0.79 | 5.60 | 0.1399 |
| **Permanent availability of PPE** |  |  |  |  |  |  |  |  |  |
| Glove |  |  |  |  |  |  |  |  |  |
| Yes | 58 (46.0) | 68 (54.0) | 126 | 1 |  |  |  |  |  |
| No | 39 (52.7) | 35 (47.3) | 74 | 1.31 | 0.3625 |  |  |  |  |
| **Facial mask** |  |  |  |  |  |  |  |  |  |
| Yes | 30 (33.0) | 61 (67.0) | 91 |  |  |  |  |  |  |
| No | 41 (37.6) | 68 (62.4) | 109 | 1.01 | 0.9694 |  |  |  |  |
| **Face shield/goggle** |  |  |  |  |  |  |  |  |  |
| Yes | 3 (50.0) | 3 (50.0) | 6 | 1 |  |  |  |  |  |
| No | 68 (35.1) | 126 (64.9) | 194 | 0.54 | 0.4576 |  |  |  |  |
| **Gown** |  |  |  |  |  |  |  |  |  |
| Yes | 33 (42.3) | 45 (57.7) | 78 | 1 |  |  |  |  |  |
| No | 38 (31.1) | 84 (68.9) | 122 | 0.62 | 0.1088 |  |  |  |  |
| **Awareness of ICC** |  |  |  |  |  |  |  |  |  |
| Yes | 24 (42.9) | 32 (57.1) | 56 | 1 |  |  |  |  |  |
| No | 2 (40.0) | 3 (60.0) | 5 | 0.89 | 0.9015 |  |  |  |  |
| I don't know | 45 (32.4) | 94 (67.6) | 139 | 0.64 | 0.1675 |  |  |  |  |
| **ICP refresher training received** |  |  |  |  |  |  |  |  |  |
| Yes | 17 (35.4) | 31 (64.6) | 48 | 1 |  |  |  |  |  |
| No | 54 (35.5) | 98 (64.5) | 152 | 1.00 | 0.9890 |  |  |  |  |
| **Poster on AEB available** |  |  |  |  |  |  |  |  |  |
| Yes | 68 (37.0) | 116 (63.0) | 184 | 1 |  |  |  |  |  |
| No | 3 (23.1) | 10 (76.9) | 13 | 0.51 | 0.3215 |  |  |  |  |
| I don't know | 0 (0.0) | 3 (100.0) | 3 | — |  |  |  |  |  |

cOR: Crude Odds Ratio; aOR: Adjusted Odds Ratio; CI: Confidence Interval, ICP: Infection Control and Prevention, ICC: Infection Control and Prevention Committee, AEB: Accidental Exposure to Body fluid; PPE; Personal Protective Equipment

Blood was the most reported body fluid involved in AEBs (87.10%).

Scissors, hollow and suture needles were the instruments involved in >80% of exposures (Figure 1).

**Figure 1.**
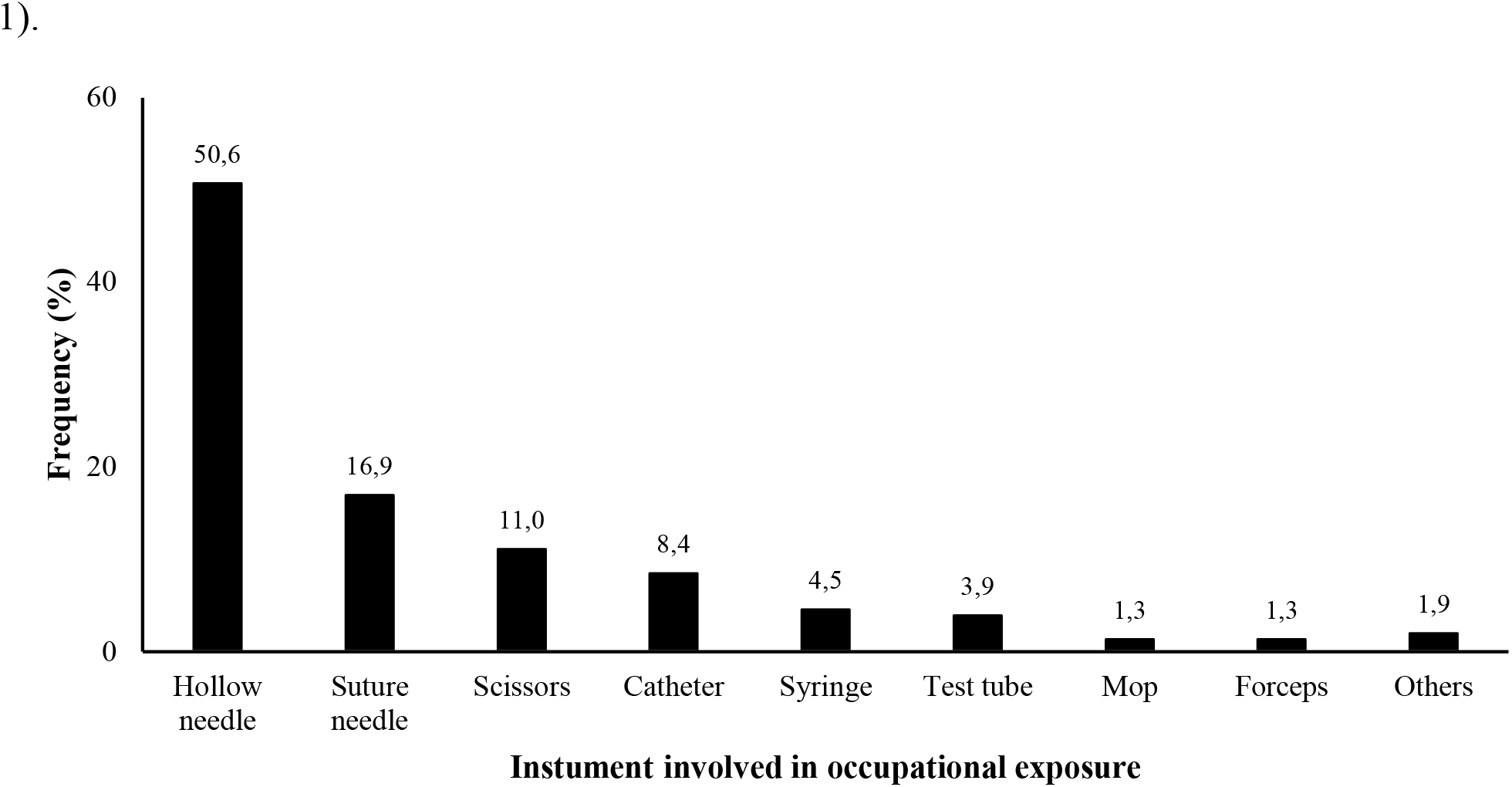
Instruments Involved in Accidental Exposures to Body Fluids among Healthcare Workers of the Buea Regional Hospital, 2023 (*n*=200)

### Circumstances of Occurrence of AEBs

Most accidental exposures (>80%) occurred while administering injections, blood sample collection, deliveries, surgery and cleaning of used equipments (Figure 2).

**Figure 2.**
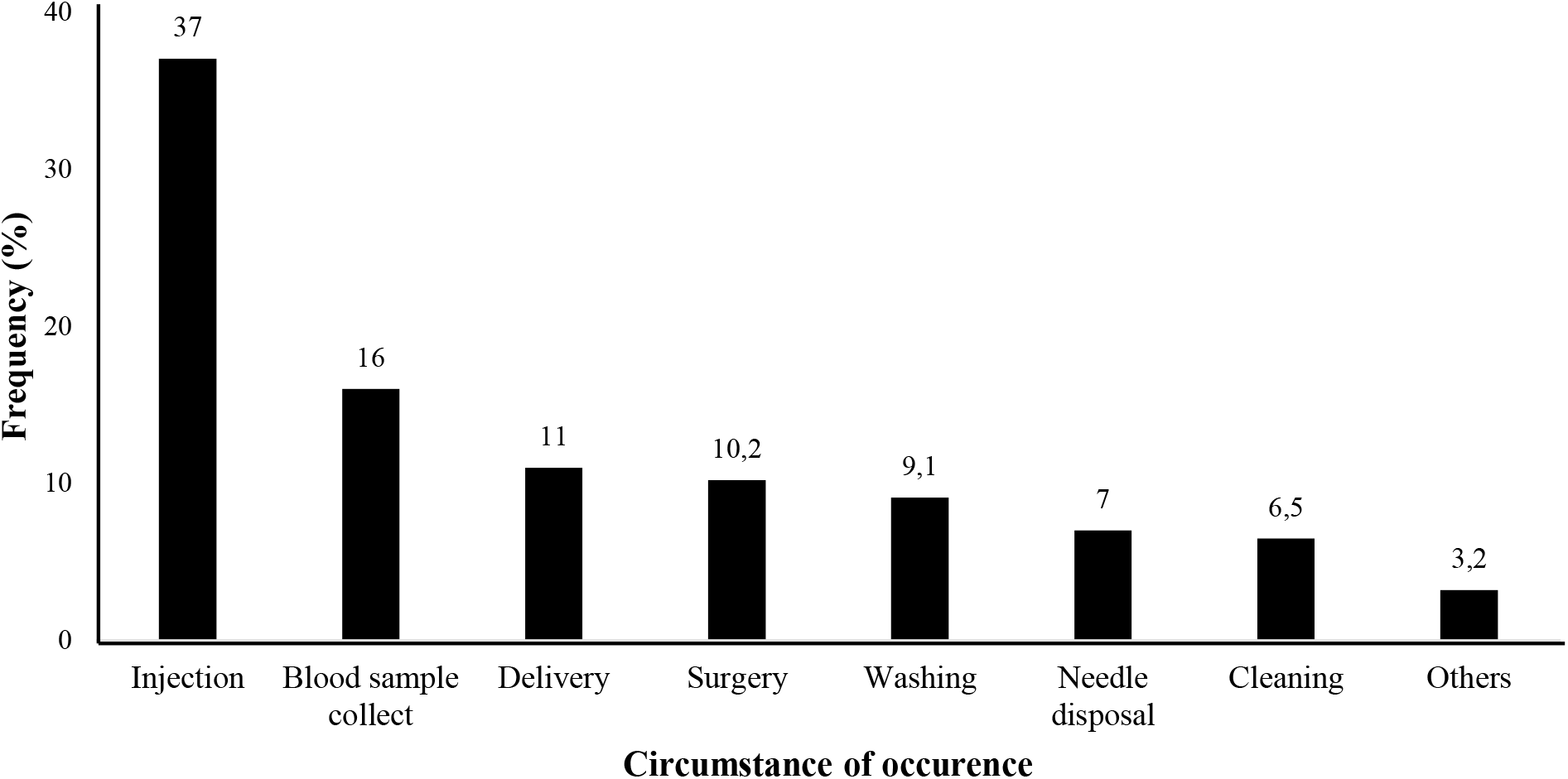
Circumstances of Occurrence of AEBs Among Healthcare Workers at the Buea Regional Hospital, 2023 (*n*=200)

### Adherence to Standard Precautions

Most of participants (54%) washed their hands after every procedure. Participants justified their non-compliance with hand-washing prescription by the tight schedule and unavailability of water (88.2%) (Figure 3).

**Figure 3.**
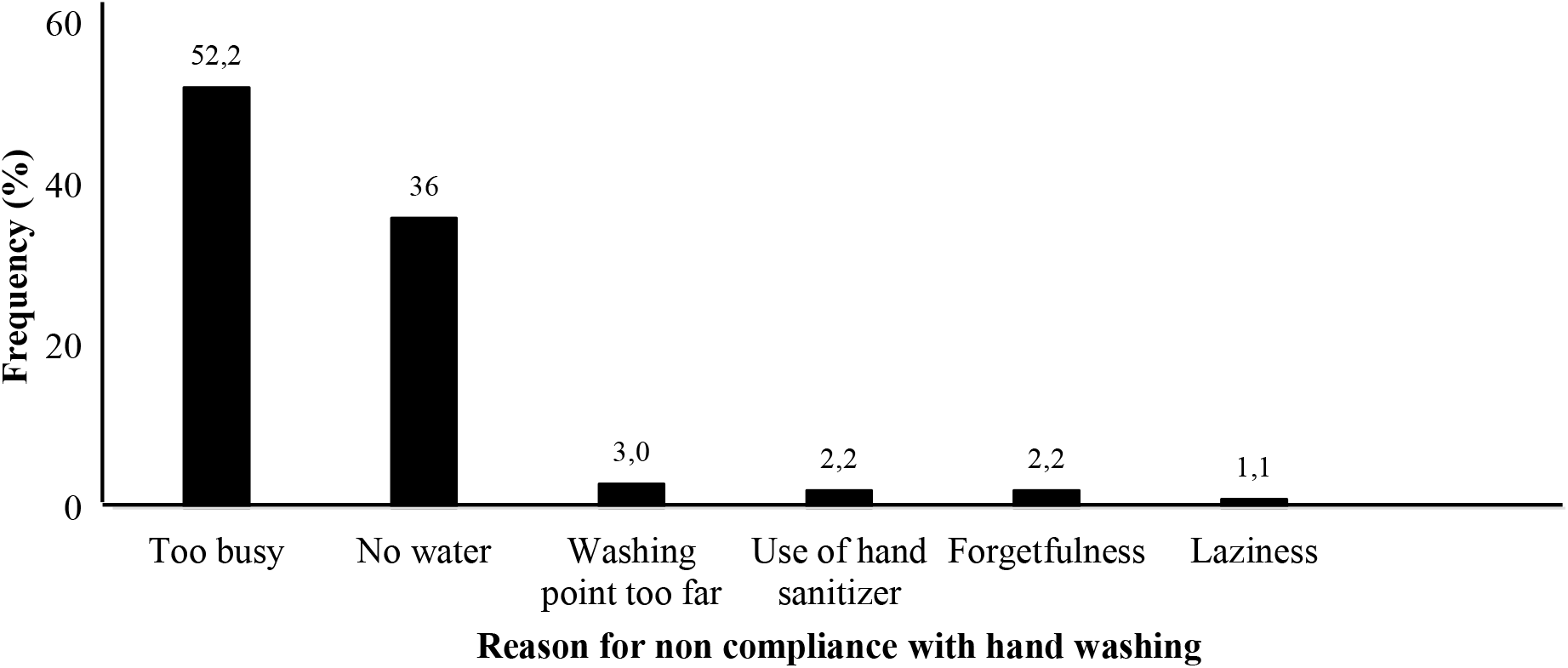
Determinants of Non-compliance with Systematic Hand washing Among Healthcare Workers at the Buea Regional Hospital, 2023 (*n*=200)

### Availability of Personal Protective Equipment (PPE)

Gloves were available in 63% of cases. Face shields and goggles were the least available PPE (3%) (Figure 4).

**Figure 4.**
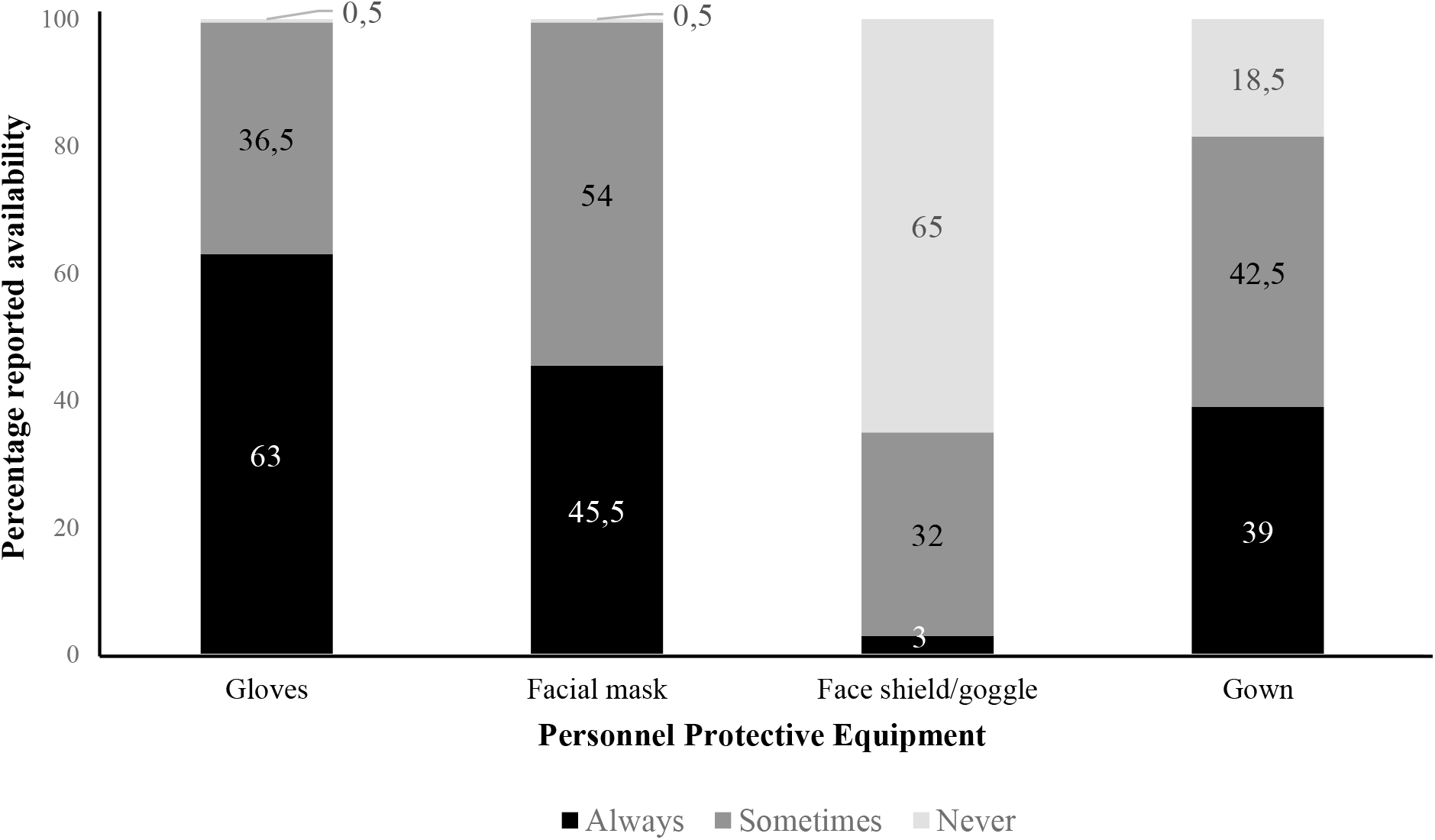
Reported Availability of PPE Among Healthcare Workers at the Buea Regional Hospital, 2023 (*n*=200)

### Reporting and post exposure Management

More than half (56.5%) of occupational exposure were not reported. Participants who had not received training on ICP (*p*-value=0.015) were more likely under declared AEB (Table 3).

**Table 3.** Factors Associated with Reporting of Occupational Exposure to Body Fluids Among Healthcare Workers at the Buea Regional Hospital, 2023 (*n*=200)

| Risk Factor | Underreporting<br>*n* (%) |  | Total<br>(100%) | cOR | *p*-value | aOR | 95%CI |  | *p*-value |
| --- | --- | --- | --- | --- | --- | --- | --- | --- | --- |
|  | Yes<br>*n*=113 | No<br>*n*=87 |  |  |  |  | Lower | Upper |  |
| **Sex** |  |  |  |  |  |  |  |  |  |
| Female | 83 (55.3) | 67 (44.7) | 150 | 1 |  |  |  |  |  |
| Male | 30 (60.0) | 20 (40.0) | 50 | 1.21 | 0.5646 |  |  |  |  |
| **Age group (years)** |  |  |  |  |  |  |  |  |  |
| 18-24 | 15 (55.6) | 12 (44.4) | 27 | 1 |  |  |  |  |  |
| 25-34 | 67 (54.5) | 56 (45.5) | 123 | 0.96 | 0.9184 |  |  |  |  |
| 35-44 | 24 (63.2) | 14 (36.8) | 38 | 1.37 | 0.5380 |  |  |  |  |
| 45 + | 7 (58.3) | 5 (41.7) | 12 | 1.12 | 0.8718 |  |  |  |  |
| **Educational level** |  |  |  |  |  |  |  |  |  |
| University | 76 (44.7) | 94 (55.3) |  | 1 |  |  |  |  |  |
| Secondary or Primary School | 11 (36.7) | 19 (63.3) |  | 1.40 | 0.4142 |  |  |  |  |
| **Unit** |  |  |  |  |  |  |  |  |  |
| Medical ward | 28 (45.2) | 34 (54.8) | 62 | 1 |  |  |  |  |  |
| Emergency | 9 (56.3) | 7 (43.8) | 16 | 1.56 | 0.4303 |  |  |  |  |
| Laboratory | 13 (72.2) | 5 (27.8) | 38 | 3.16 | **0.0493** |  |  |  |  |
| Surgical ward | 23 (59.0) | 16 (41.0) | 39 | 1.75 | 0.1781 |  |  |  |  |
| Obstetrics and gynecology | 12 (54.5) | 10 (45.5) | 22 | 1.46 | 0.4501 |  |  |  |  |
| Pediatrics | 6 (50.0) | 6 (50.0) | 12 | 1.21 | 0.7584 |  |  |  |  |
| Dental service | 6 (100.0) | 0 (0.0) | 6 | 0.00 | 0.9863 |  |  |  |  |
| Intensive care unit | 3 (50.0) | 3 (50.0) | 8 | 1.21 | 0.8205 |  |  |  |  |
| Out - patient consultation | 13 (68.4) | 6 (31.6) | 19 | 2.63 | 0.0817 |  |  |  |  |
| **Professional status** |  |  |  |  |  |  |  |  |  |
| Assistant nurse & Cleaner | 10 (50.0) | 10 (50.0) | 20 | 1 |  |  |  |  |  |
| Laboratory technician | 12 (70.6) | 5 (29.4) | 17 | 2.40 | 0.2079 |  |  |  |  |
| Physician | 9 (47.4) | 10 (52.6) | 19 | 0.90 | 0.8695 |  |  |  |  |
| Midwife & Nurse | 82 (56.9) | 62 (43.1) | 144 | 1.32 | 0.5585 |  |  |  |  |
| **Years of experience 1** |  |  |  |  |  |  |  |  |  |
| <5 | 74 (56.5) | 57 (43.5) | 131 | 1 |  |  |  |  |  |
| 5+ | 39 (56.5) | 30 (43.5) | 69 | 1.00 | 0.9964 |  |  |  |  |
| **Years of experience 2** |  |  |  |  |  |  |  |  |  |
| <10 | 90 (53.9) | 77 (46.1) | 167 | 1 |  | 1 | — | — |  |
| 10+ | 23 (69.7) | 10 (30.3) | 33 | 1.97 | 0.0982 | 2.18 | 0.99 | 5.12 | 0.0603 |
| **ICC available** |  |  |  |  |  |  |  |  |  |
| Yes | 36 (64.3) | 20 (35.7) | 56 | 1 |  |  |  |  |  |
| No | 3 (60.0) | 2 (40.0) | 5 | 0.83 | 0.8485 |  |  |  |  |
| I don't know | 74 (53.2) | 65 (46.8) | 139 | 0.63 | 0.1607 |  |  |  |  |
| **ICP refresher training received** |  |  |  |  |  |  |  |  |  |
| Yes | 34 (70.8) | 14 (29.2) | 152 | 1 |  | 1 | — | — |  |
| No | 79 (52.0) | 73 (48.0) | 48 | 2.22 | **0.0234** | 2.41 | 1.21 | 5.01 | **0.0150** |
| **Poster on AEB available** |  |  |  |  |  |  |  |  |  |
| Yes | 101 (54.9) | 83 (45.1) | 184 | 1 |  |  |  |  |  |
| No | 10 (76.9) | 3 (23.1) | 13 | 2.74 | 0.1353 |  |  |  |  |
| I don't know | 2 (66.7) | 1 (33.3) | 3 | 1.64 | 0.6871 |  |  |  |  |

cOR: Crude Odds Ratio; aOR: Adjusted Odds Ratio; CI: Confidence Interval ICP: Infection Control and Prevention, ICC: Infection Control and Prevention Committee, AEB: Accidental Exposure to Body fluids

The main reasons for not reporting exposure were lack of awareness of the reporting system (44.4%) and the perception that the risk of contamination was low (30.3%). Most participants (76.0%) were of the opinion that there was a post-exposure unit for case management, where PEP (post exposure prophylaxis could be administered (Figure 5).

**Figure 5.**
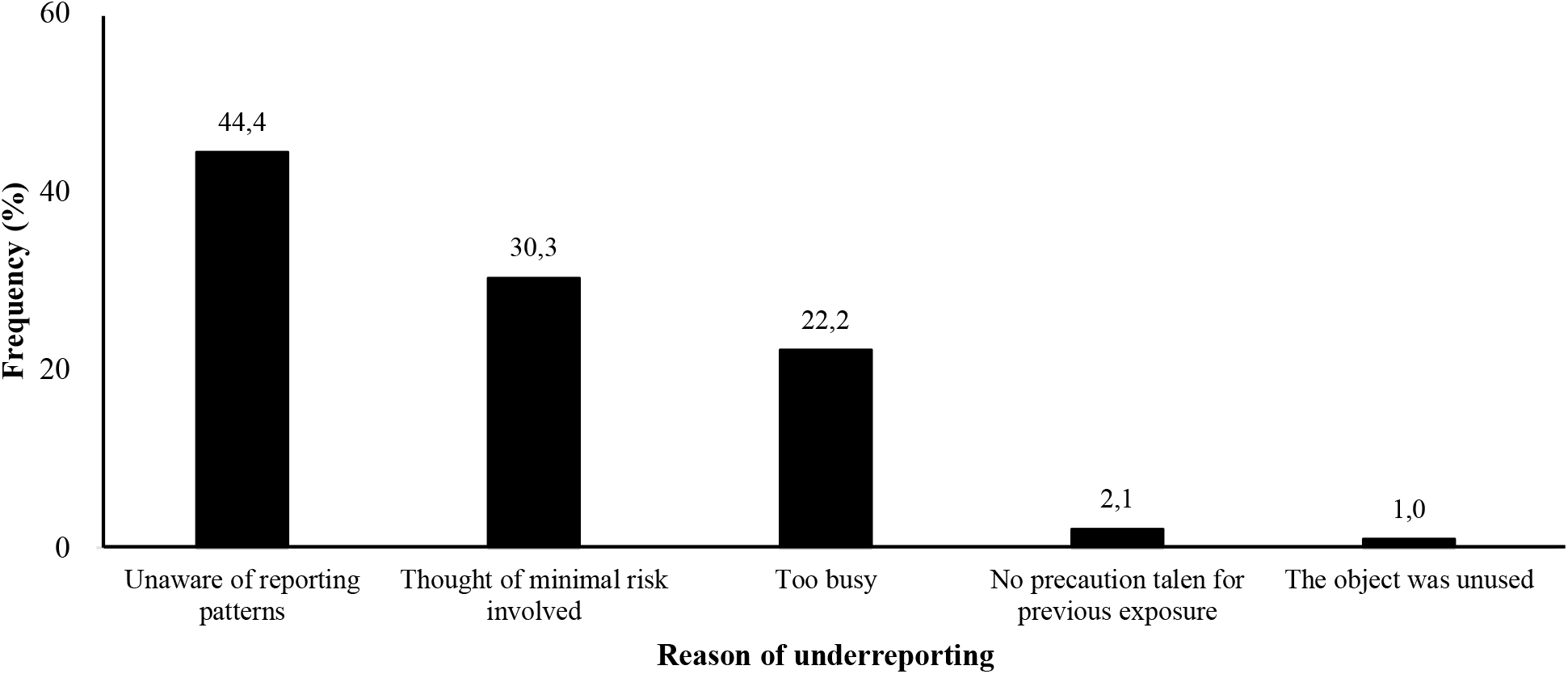
Reported Availability of Personal Protective Equipment (PPE) Among Healthcare Workers at the Buea Regional Hospital, 2023 (*n*=113)

Participants who experienced exposure washed the affected area with soap and water (52.8%) and 4% took a course of post-exposure prophylaxis.

### Observance of Post-exposure Management Procedures

Most participants (92%) reported that post-exposure management posters were available in clinical departments. The rules guiding ICP team membership could not be enumerated by participants. Such structures were not fully functional. Cases of exposure were directed to the vaccination unit who occasionally conducted sensitization on occupational exposures within the hospital. Furthermore, there was no registry for accidental exposures to BBFs.

## Discussion

The rather high prevalence of AEB (83.5%) reported in the present investigations is consistent with the global situation of vulnerability among HCW [31] and the trends observed in Yaoundé District Hospitals were more than 50% of HCWs had experienced AEB over a one-year period [29].

The study was conducted in Buea, where several armed groups steer security challenges since 2016 in the two historically Anglophone regions of Cameroon [20]. Social unrest and conflicts are major constrains to the performance of the healthcare system, including the quality of services delivered, as attacks on HCWs and destruction of health facilities. Common problems such as lockdowns disrupt hospital-based activities exacerbating the burden on an already fragile healthcare system. The perception of insecurity and uncertainty is an impediment to HCWs working environment leading to physical and mental fatigue. The pre-existing problem of HCWs shortage worsen the situation [32] leading to an increase in the incidence of AEB [27,29,36,37].

Most AEB occurred through NSIs (48.5%) and splashes (35.5%), corroborating findings in Uganda (55%) [38]. However, disparities in the triggers of exposures were reported in Cameroon and Ethiopia [26,29,39]. The high rate of exposures through NSIs and splashes call into question the effectiveness of training on infection prevention and control, highlighting the need to strengthen educational activities, the need to scale up the supply of PPE and adherence to standard precautions.

HCWs from medical (aOR=5.95) and pediatric (aOR=10.9) services were more likely to experience NSI. settings findings differ from observations in other district level settings in Cameroon. Such differences may indicate varied approaches to the implementation and enforcement of IPC measures [27].

Females were 2.9 times more likely to experience a splash exposure than male HCWs. Exposure was higher among HCW operating in the obstetric unit. Therein, the splashes involved the amniotic fluids which were described as being common in the delivery room [39].

Standard precautions are designed to reduce the risk of transmission of bloodborne pathogens. They are a minimum set of precautions in the delivery of care to all patients. Key precautions include hand washing, use of personal protective equipment, respiratory etiquette, prevention of needlestick and sharp injuries [43,44]. Ideally, universal precautions are followed by all workers. However, the present study revealed that only 54% of HCWs systematically washed their hands after a health care procedure. The most common reasons for poor adherence to hand washing were a busy schedule and unavailability of water. Hand hygiene is a major component of standard precautions and the most effective method of preventing the transmission of hand-associated pathogens [36,45,46]. Failure to follow this measure increases the spread of infectious agents, including life threatening pathogens such as hepatitis B [47].

Recapping used needles was practiced by over 85% of HCWs. Recapping needle is a risky practice that often results in the accidental puncture of the fingers or hand, leading to exposure to hazardous chemicals, drugs or infectious agents. Even though needle recapping is not recommended, it the scoop-cap method of placing the cap on a hard surface and using one hand for recapping is advised when necessary in resources-limited settings where equipment with safety shield is not available [48].

Gloves were always available only in 63% of cases and face shields/goggles were the least available PPE. This increases the risk of splash into eye, nose or mouth [49]. The availability of hand-washing facilities and the provision of PPE are measures aimed at creating a safe environment for patients and caregivers. Sub-optimal adherence to universal precautions may reflect flaws in supervision at intermediate and central levels. In that regards, emphasis on the role of the ICC, internal and external monitoring guidelines is needed to promote compliance [36].

Less than a third of HCWs (24%) had received training in infection control and prevention. Training with updating is associated with improved knowledge, positive changes in attitudes and practices and a reduction of occupational exposure.

Most exposures occurred during the administration of injections, blood collection and delivery, involving hollow and suture needles. Most exposures occurred while performing surgical procedures, recapping needle and in the disposal of used syringes [50,51]. Other factors favoring exposure include handling an uncooperative patient, the long working hours…[52].

More than half of the exposures were not declared (56.5%) and HCWs who had not received refresher training on ICP were most likely to avoid reporting. This can be explained by the lack of knowledge of the reporting channels and the underestimation of the health risks associated with exposure [28,53]. There is a need to involve all cadres in the training, implementation, monitoring and evaluation of ICP measures. Underreporting may be related to the fact that the ICC at the Buea Regional Hospital was scantily known to HCWs. In addition, there was no dedicated registry for AEBs [26,53].

Despite the low level of reporting, two-thirds (67.2%) of those who experienced AEB received post-exposure prophylaxis. Most of the HCW exposed knew about the management requirements, and proceeded with emergency post-exposure treatment without filing the notification [28].

## Conclusions

Most HCW of the Buea Regional Hospital had experienced an accidental exposure to blood and body fluids over the last year. Accidental exposure to body fluids occurred in a context of poor adherence to universal precautions resulting from an inadequate supply of Personal Protective Equipment (PPE) and inadequate hand washing facilities. There is an urgent need to establish an effective Infection Control & Prevention team to pilot educational activities, monitoring and management system for the prevention of AEB.

## Data Availability

All data produced in the present work are contained in the manuscript

## Abbreviations

AEB: Accidental Exposure to Body fluids
AIC: Akaike Information Criterion
aOR: Adjusted Odds Ratio
BBF: Blood and Body Fluid
CI: Confidence Interval
cOR: Crude Odds Ratio
HBV: Hepatitis B Virus
HCW: Healthcare Worker
HIV/AIDS: Human Immunodeficiency Virus/Acquired Immunodeficiency Syndrome
HVC: Hepatitis C Virus
ICC: Infection Control and Prevention Committee
ICP: Infection Control and Prevention
NSI: Needlestick and Sharp Injury
PPE: Personal Protective Equipment

## Declaration

### Authors’ Contribution

Drafting of the study protocol, data collection, analysis and interpretation: B.A.C & F.Z.L.C; Drafting, data analysis and editing of manuscript: N.D.L & F.Z.L.C.; Critical revision of protocol and manuscript: F.Z.L.C and I.T.; Conception, design and supervision of research protocol and implementation, data analysis plan, revision, editing and final validation of the manuscript: I.T.

### Ethical Approval Statement

The protocol was approved by Institutional Review Board (IRB) of the Faculty of Medicine and Biomedical Sciences of Yaoundé and the ethical clearance: N°0344/UY1/FMSB/VDRC/DAASR/CSD granted. Informed consent was obtained from participants prior to inclusion in the study. All methods were performed according to relevant guidelines and regulations.

### Consent for publication

Not applicable.

### Availability of data and materials

All data generated or analyzed during this study are included in this published article.

## Acknowledgements

Our gratitude goes to the health staff who agreed to participate in this study and to the manager of the Buea Regional Hospital who gave an authorization to conduct the study.

